# Cohort Profile: Health and Demographic Surveillance System in Mirzapur, Tangail, Bangladesh

**DOI:** 10.1101/2025.07.21.25331957

**Authors:** Senjuti Saha, Naito Kanon, Mohammad Shahidul Islam, Mohammad Shameem Hassan, Yogesh Hooda, Lubana Tanvia, Qazi Sadequr Rahman, Rajib C Das, Shampa Saha, Sanwarul Bari, AKM Tanvir Hossain, Md Shariful Islam, Gary L Darmstadt, Shams El Arifeen, Samir K Saha

## Abstract

The Mirzapur Health and Demographic Surveillance System (MHDSS), established in 2007 in Tangail, Bangladesh, systematically monitors population and health dynamics across 10 unions, covering 316,628 individuals as of 2022. It collects longitudinal data on vital events, including births, deaths, migrations, and immunizations, every four months, with socio-economic updates every five years. Unique to the MHDSS is its integration of an Infectious Disease Surveillance (IDS) program, which evaluates the burden of invasive diseases in under-five year old children every week through community, hospital, and laboratory data linkage.

The surveillance area spans predominantly rural terrain with a subtropical climate and includes good healthcare infrastructure, anchored by the Kumudini Women’s Medical College and Hospital. Data collection employs an Android-based system for real-time updates, ensuring high-quality and accessible data. Over the years, MHDSS has generated pivotal findings, including declining birth rates, and reduced neonatal and child mortality. It has facilitated research on newborn care, vaccine impacts, respiratory diseases, and environmental health.

Data is available for collaborative research through CHRF’s Data Center upon request. Future plans include expanding analyses of non-communicable diseases, mental health, and the impacts of urbanization and climate change on health outcomes.

**Key Features:**

⍰ The Mirzapur Health and Demographic Surveillance System (MHDSS) was established in 2007 to continuously monitor population and health dynamics.
⍰ As of 2022, it covers a population of 316,628 individuals across 81,296 households, with 99% participation rate.
⍰ Vital events such as pregnancies, births, deaths, migrations, and immunizations of children under five, are collected every four months, while socio-economic data are updated every five years by trained female data collectors.
⍰ The Infectious Disease Surveillance program embedded within MHDSS evaluates trends in infectious diseases and impacts of immunizations in children under five years of age and integrates community health data with hospital and laboratory findings for comprehensive analysis.
⍰ Cohort data are accessible for collaborative research via the Child Health Research Foundation Data Center, subject to project description and approval.

## Why was the cohort set up?

In low-and-middle income countries like Bangladesh, limited administrative and health infrastructure preclude continuous monitoring of health indicators at the community level (1). Nationally representative Bangladesh Demographic and Health Surveys (BDHS) are conducted to monitor vital statistics and important health and demographic indicators. Since BDHSs are conducted every 3-4 years, the surveys overlook short-term changes and seasonal variations in health data (2). BDHSs are also limited by the coverage of broader groups of communicable and non-communicable diseases (3) and long intervals between the surveys often cause recall bias and fail to capture the immediate impact of health interventions. While such surveys provide overall national estimates, they may not capture specific community-level dynamics, and are insufficiently powered to support intersectional analyses (4,5). To fill this data gap and foster innovative community-level interventions, a surveillance system was established in Mirzapur sub-district of Bangladesh in 2007.

The Mirzapur Health and Demographic Surveillance System (MHDSS) was built to collect community-specific data at short intervals and track population fluctuations and swiftly detect health threats, including infectious disease trends, patterns and outbreaks and environmental hazards. Mirzapur was an ideal location for community surveillance because if harbors a large philanthropic hospital, Kumudini Women’s Medical College and Hospital (KWMCH), in the sub-district. The MHDSS was initiated with the vision to generate baseline data to monitor demographic and health related events such as births, deaths, migrations, pregnancy outcomes, immunization of children under five years of age and causes of death of the Mirzapur community.

## Where is the HDSS area?

The HDSS is in Mirzapur, a sub-district of Tangail district under the Dhaka division. It is positioned about 67km north of Dhaka. Mirzapur has a subtropical climate and, during the monsoon season (July-October), the area encounters substantial rainfall and flooding. The lifestyle in Mirzapur is predominantly rural, with farming as the primary source of employment.

In recent years, there has been a growing presence of small-scale industries and businesses in Mirzapur. As of 2024, the population of Mirzapur included approximately 423,708 individuals. The MHDSS operates in 10 unions that include 156 villages, representing a population of 316,628 living in 81,296 households. The MHDSS area includes several healthcare facilities. The government-run Mirzapur Upazila Health Complex, which is a 31-bed primary hospital offering essential outpatient care, immunizations, and delivery services, and union-level health and family welfare centers provide primary healthcare services to the local population. Mirzapur also has approximately 35 non-profit government-run community clinics and approximately 18 private clinics. The most prominent health facility in this area is KWMCH with 1,050 hospital beds. KWMCH offers a wide range of medical services including gynecology, obstetrics, pediatrics, surgery and internal and emergency medicine, and plays a crucial role in providing high-quality and accessible healthcare in Mirzapur.

## Who is in the cohort?

The MHDSS covers all residents in the 10 unions of Mirzapur (Figure 1a). Unions have been sub-divided into 19 clusters, each containing 80 blocks. On an average, each block serves 50 households, bringing the total number of households served by the 19 clusters to ∼4,000.

**Figure 1.**
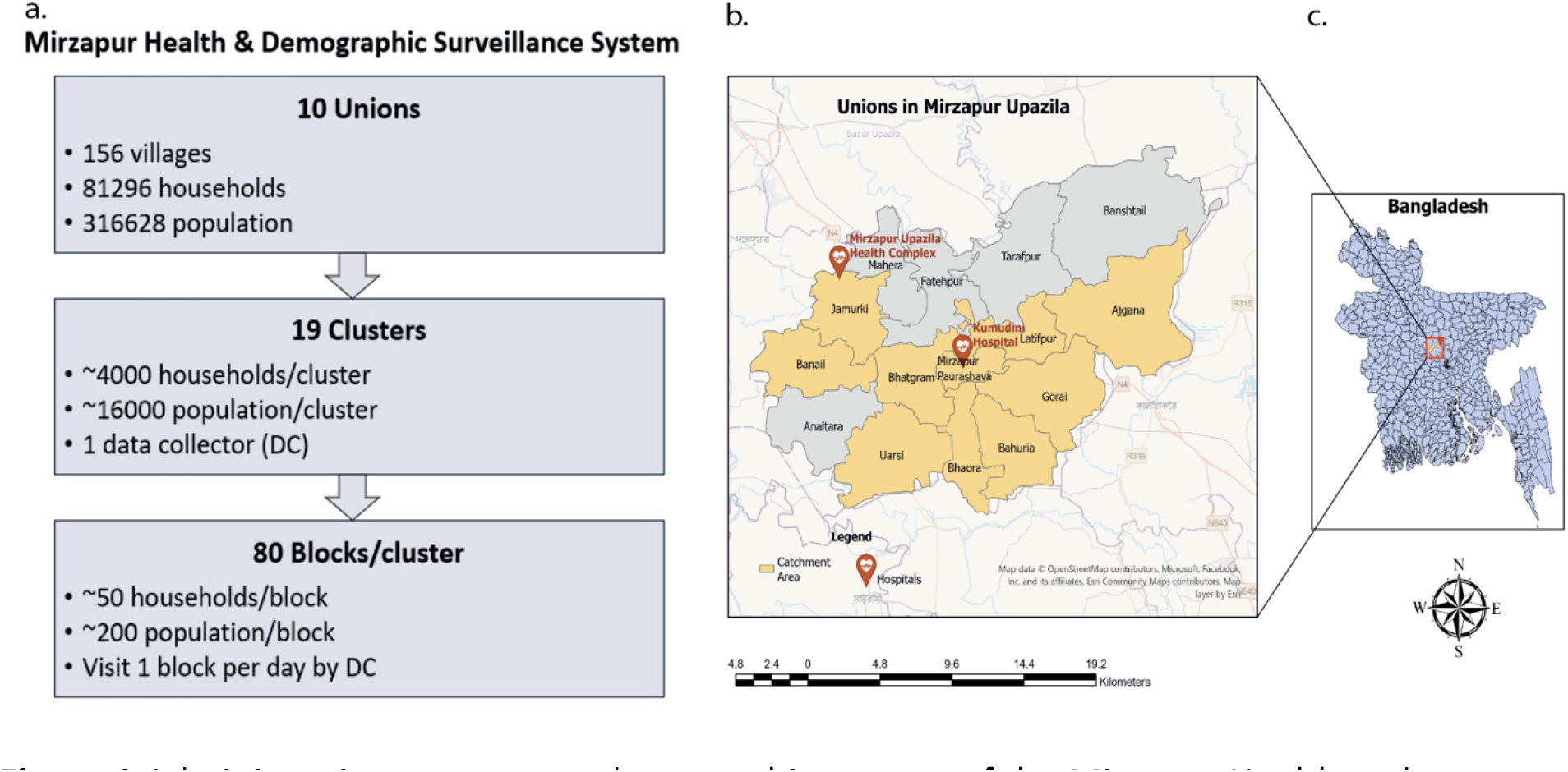
Administrative structure and geographic context of the Mirzapur Health and Demographic Surveillance System (MHDSS) in Bangladesh. (a) Sub-division of unions, clusters and blocks in the MHDSS; (b) Map of the MHDSS area with the 10 unions covered by surveillance highlighted in yellow; (c) Map of Bangladesh with a red box indicating the location of Mirzapur. **Alt text:** Diagram of the Mirzapur Health and Demographic Surveillance System administrative structure with 10 unions subdivided into 19 clusters of 80 blocks, the MHDSS area map highlighting Surveillance unions in yellow, and a map of Bangladesh with a red box marking the location of Mirzapur.

A household is defined as a unit consisting of individuals who live under the same roof and share food from the same kitchen. A resident is a person who stays in a household in the MHDSS area continuously for at least four months a year or lives outside the MHDSS area but returns at least once a month to stay overnight.

Each household in the MHDSS has a unique nine-digit household ID, which includes a three-digit Village Code, a four-digit “Bari” Code, and a two-digit Household Code. A trained data collector (DC) visits approximately 50-60 households per block daily to record information from household members. For new household or member registrations, a DC informs the household head about the purpose and benefits of the surveillance, obtains consent, and ensures confidentiality. Informed consent is recorded via signature or thumb impression and remains valid until withdrawal.

Each resident is assigned a unique 11-digit Current Identification Number (CID) during registration. The CID consists of a two-digit individual number appended to the household ID, starting from 01 for the household head. Additionally, each household member receives a unique eight-digit Permanent Identification Number (PID), comprised of the three-digit Village Code and a five-digit Individual Number. The CID changes with internal migration or household splits, while the PID remains unchanged unless a resident migrates out of the MHDSS area.

In households with children under five years of age, trained DCs provide a child card for free healthcare access at KWMCH. Pregnant women receive a mother card to track their pregnancies and post-childbirth movements.

Residents enter the surveillance population through registration, birth, in-migration (permanent move into the MHDSS area), internal in-migration (moving within the MHDSS area), or split-in (household splits). Residents are excluded due to consent withdrawal, death, out-migration (permanent move out of the MHDSS area), or household demolition.

### Nested Invasive Disease Surveillance (IDS)

IDS began in 2009 with support from the World Health Organization (WHO) to assess burden of infectious diseases among under-five children. Currently, IDS covers four of the ten unions (Figure 1b) divided into 60 clusters and five blocks per cluster, covering about 16,000 under-five children per cluster. Village health workers (VHWs) assess every under-five child weekly. If the caregiver reports a child to be sick, the VHW assesses whether the child matches the invasive disease criteria using an algorithm developed following WHO guidelines (7), and refers the child according to the guidelines to KWMCH. Children referred by VHWs or brought in by their caregivers to KWMCH are screened by physicians. During the screening of children, physicians record demographic information including the clinical signs, symptoms and diagnosis. Data on hospital investigations, antibiotic history and vaccinations are recorded. Children admitted to KWMCH are followed up, and their outcome and diagnoses are recorded at the time of discharge.

## How often have they been followed up?

Overall, the surveillance participation rate during the four-month follow-up is 100%. Vaccine coverage in this population is high, with at least 97% coverage for at least one EPI vaccine among under-five children. Each data collection round takes four to five months, updating demographic events and socio-economic information as needed; socioeconomic data is fully updated every 5 years.

## What has been measured?

The surveillance area, including boundaries and GPS coordinates of households, was mapped at the onset of the HDSS and is regularly updated as new households enter the database. Information collected for the surveillance is presented in Table 1.

**Table 1.** Information collected in the Mirzapur Health Demographic Surveillance System and the nested Infectious Diseases Surveillance System.

| Index | Frequency of follow-up | Information |
| --- | --- | --- |
| **Individuals** | Every 4 months | Name, sex, date of birth, religion, relation with the household head, educational attainment, occupation, marital status, number of spouses, contact number |
| **Pregnancy** | Every 4 months | Name, date of last menstrual period (LMP), pregnancy status, delivery outcome (live birth, still birth, miscarriage), delivery place, number and type of miscarriage (induced, spontaneous) |
| **Births** | Every 4 months | Name of newborn, sex, date and place of birth, type of birth attendant |
| **Deaths** | Every 4 months | Name of deceased, date and place of death, cause of death through verbal autopsy, hospitalization of the deceased before 14 days of death, place of cremation/ burial. |
| **Migrations** | Every 4 months | Name of migrant, type of migration, date and cause of migration, destination after migration (if possible) |
| **Immunizations** | Every 4 months | Type of vaccine, date of vaccination, vaccination center |
| **Socio-economic status** | Every 5 years | Household construction type, roof materials, rooms, facility locations, house ownership, rent, drinking water sources, hand-washing facilities, latrine type, electricity, cooking energy, ownership of household equipment, livestock, land, relatives abroad, NGO membership |
| **Infectious Disease Surveillance (IDS)** | Every week<br>(in 4 of 10 unions) | Symptoms of invasive diseases (pneumonia, severe pneumonia, meningitis, very severe disease, typhoid fever), referrals, diagnosis, treatment outcomes |

During household registration, DCs record details such as electricity availability, drinking water source, and toilet facilities, along with socio-economic status indicators like asset and animal ownership, land ownership, overseas family members, and NGO membership. Demographic indicators related to mortality, migration, marriage, fertility of ever-married women, and immunization of under-five children are also recorded. In addition, information on causes of death, hospitalization before death, and burial/cremation details are collected through verbal autopsy. IDS data are linked with hospital data using the CID/PID and referral slip numbers.

The data collection team includes field workers (FWs), DCs, field supervisors (FSs), a senior field research officer (SFRO), and a field research manager (FRM). DCs collect and update data, FWs update socio-economic status and assist as backup DCs, and FSs monitor DCs and conduct quality assessments. Any data inconsistencies are corrected after discussions in weekly meetings. The SFRO supervises daily activities, prepares monthly plans, and generates child and mother cards. The FRM coordinates field studies and maintains a migration tracking form. Meetings are held at the end of each round of data collection to address data collection challenges and provide refresher training.

From 2007-2013, data were collected using paper forms, but since 2013, an Android-based customized data collection tool is used. The database is maintained on a Microsoft SQL Server, hosted by the CHRF Data Center. Data synchronization is conducted daily to ensure real-time availability of data, and data are backed up weekly. Access to the database is provided via ODBC with read-only permission for data exporting.

## What has it found?

Demographic indicators of the MHDSS from 2007 to 2022, analyzed at five-year intervals, are presented in Table 2.

**Table 2.**
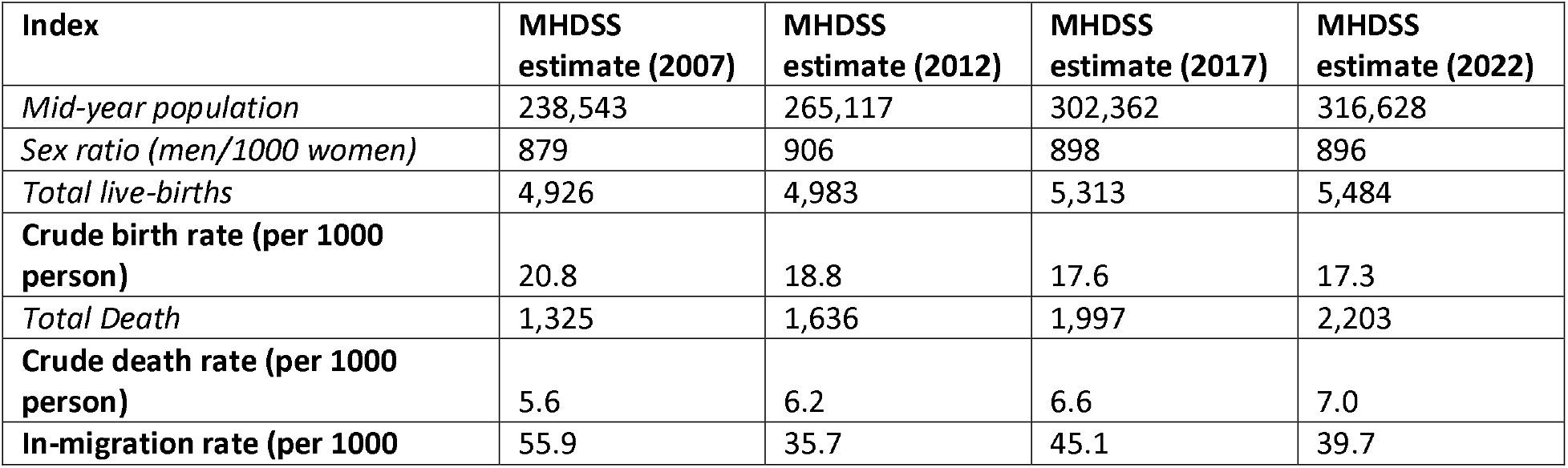
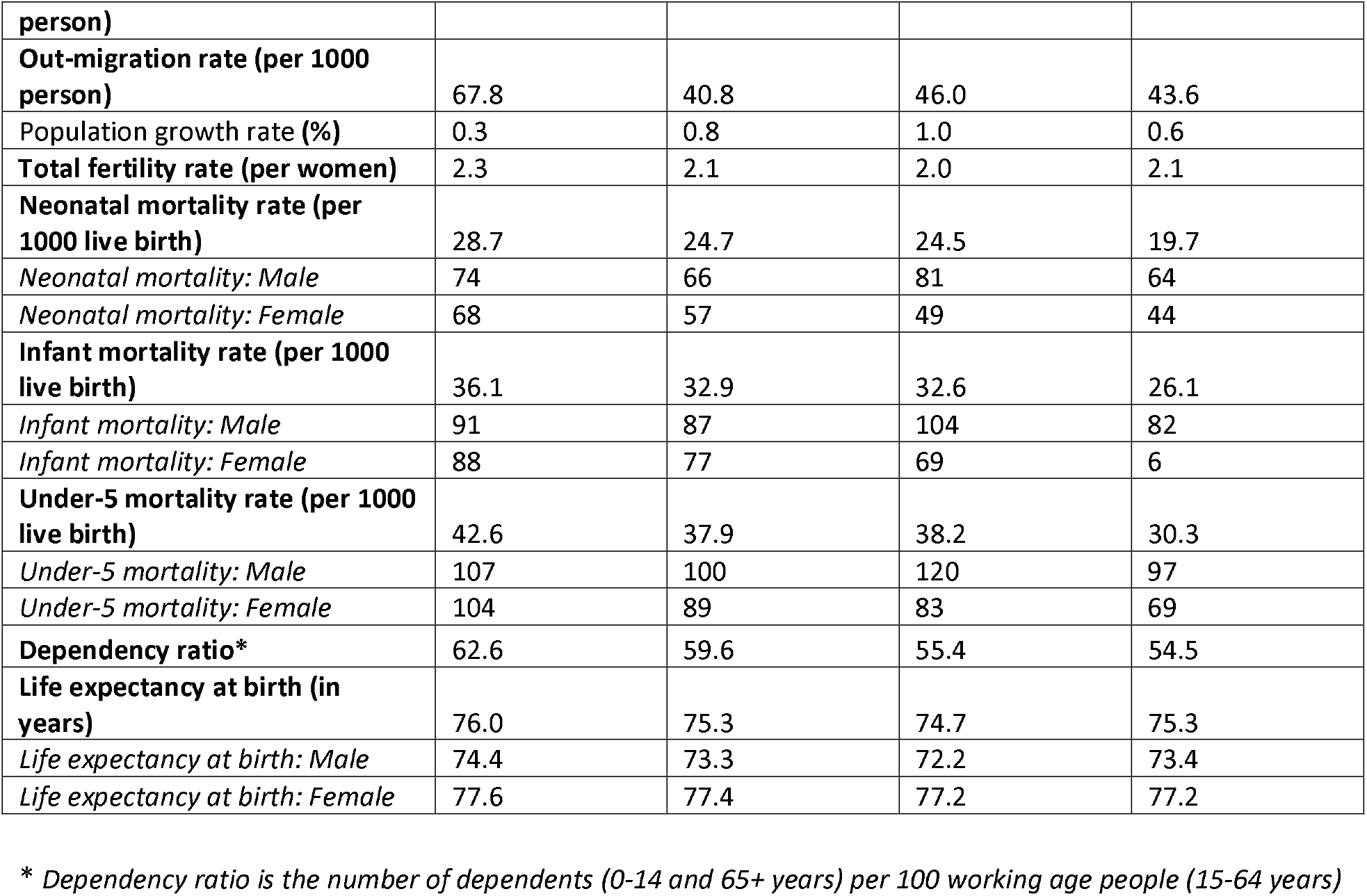
Demographic characteristics of Mirzapur Health Demographic Surveillance System.

### Changes in population structure

The population of the MHDSS increased from 238,543 in 2007 to 316,628 in 2022 (Figure 2). There was a shift in male-female ratio from 0.879 (i.e. 879 men per 1000 women) in 2007 to 0.896 in 2022, likely due to higher out-migration among men compared to women. A large proportion of men aged 20-34 migrate out of Mirzapur for jobs or education, while women aged 15-24 migrate in upon marriage.

**Figure 2.**
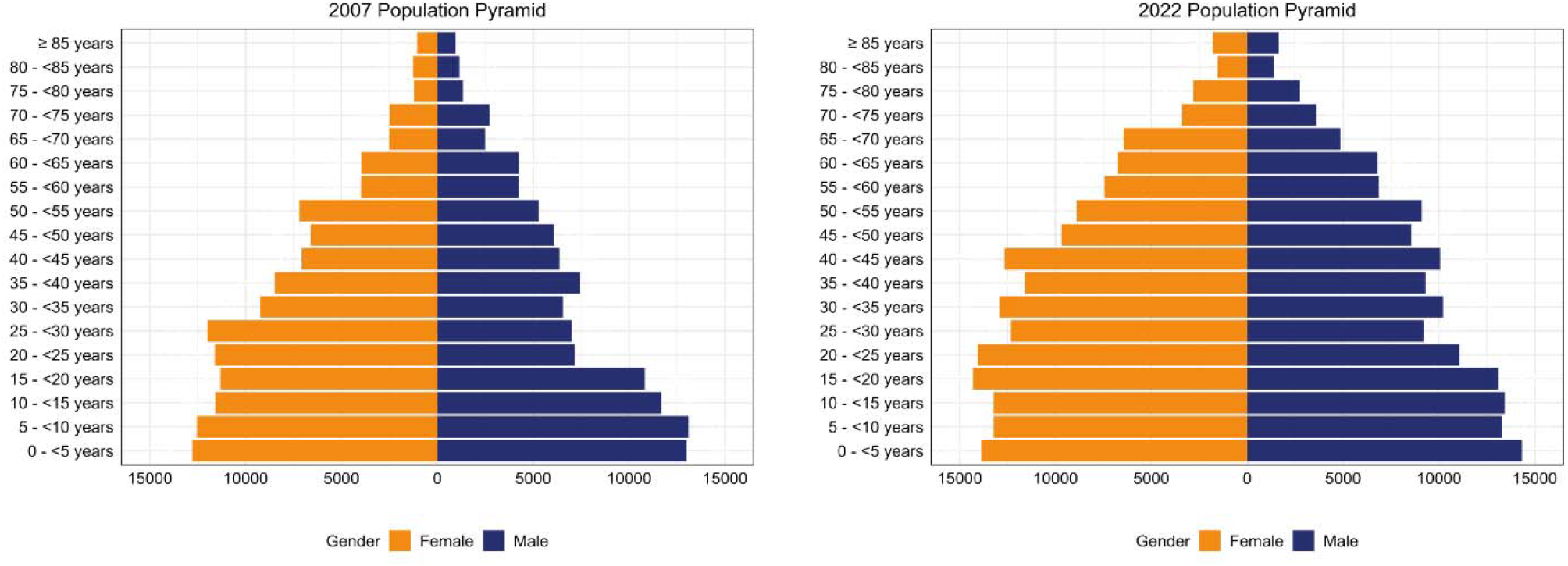
Age distribution of the Mirzapur Health and Demographic Surveillance System population in 2007 and 2022. **Alt text:** Two population pyramids comparing age and sex structure in the Mirzapur HDSS area for 2007 and 2022, with male population bars extending left and female bars extending right across 5 year age groups, showing a wider youth base in 2007 and a narrower base with more evenly distributed adult age groups in 2022.

### Rate of Natural Increase

The MHDSS data showed a decline in crude birth rate from 20.8 per thousand in 2007 to 17.3 by 2022 (Table 2), in comparison to the national estimates by the World Bank (8), which showed a steeper decline from 23.9 in 2007 to 17.5 in 2022. The crude death rate at MHDSS was 5.6 per thousand population in 2007 and 7.0 in 2022, similar to the estimates in the national data (9) - 6.8 in 2007 and 5.3 in 2022 (Table 2). Consequently, the rate of natural increase at the MHDSS steadily decreased from 15.2 per thousand in 2007 to 10.3 in 2022, more rapidly than national estimates that decreased from 17.2 to 12.2 over the same period (Table 2).

### Trends in Child Mortality

In the MHDSS, neonatal mortality decreased from 28.7 per thousand live births in 2007 to 19.7 in 2022, less rapidly than the decline in the national estimates from 33.3 in 2007, to 17.4 in 2022 (10) (Table 2). Under-5 mortality in the MHDSS reduced from 42.6 per thousand live births in 2007 to 30.3 in 2022, showing less rapid decline than the national estimates of 57.7 in 2007 to 28.8 in 2022 (11) (Table 2).

### Research output

Since its establishment, the Mirzapur HDSS has been a highly productive research platform, generating critical insights into a wide range of public health challenges. It has facilitated large-scale epidemiological studies on vaccine impact, infectious diseases, respiratory illnesses, and environmental health, leading to significant contributions to global and national public health policies (Table 3).

**Table 3.** Research studies conducted in Mirzapur HDSS.

| Study title | Objective of study | Population under study | Donor | Year of Completion | Reference |
| --- | --- | --- | --- | --- | --- |
| Pneumococcal Vaccines Accelerated Development and Introduction Plan (PneumoADIP) | To provide information on invasive pneumococcal disease incidence, seasonal variations, drug-resistance patterns, and serotype composition associated with community-acquired disease | Children aged 1-59 months | GAVI Alliance | 2007 | (12) |
| Group B Streptococcus (GBS) among Pregnant Women and Newborns | To examine the maternal GBS carriage rate and determine the magnitude of GBS transmission from mother to baby | Pregnant women and their newborns | Centers for Disease Control and Prevention | 2013 | (13) |
| Vaccination and the Paediatric Microbiome | To define the nasopharyngeal microbiome in newborns by monthly NP swabs during their first year after birth | Newborns aged 5-7 days, their mothers and immediate siblings | University of Washington | 2016 | In preparation |
| Impact of Pneumococcal Conjugate Vaccine (PCV) on Nasopharyngeal Carriage in Mirzapur, Tangail | To compare the prevalence of pneumococcal carriage among vaccine-eligible and non-eligible age groups before and after the introduction of PCV-10 | Children aged 4-35 months and their mothers | IHME | 2016 | In preparation |
| Group B Streptococcus colonization in mother-newborn dyads and association with serotype-specific capsular antibodies in low and middle income South Asian and African countries | To determine the prevalence of GBS rectovaginal colonisation and serotype distribution among pregnant women at the time of delivery in African and South Asian low- and middle-income countries using standardised methods | Pregnant mother and their newborns | South Africa Medical Research Council Vaccines and Infectious Diseases Analytics Research Unit | 2017 | (14) |
| Nasopharyngeal Pneumococcal Carriage Study in South Asian Infants | To estimate the prevalence of pneumococcal carriage and serotype distribution among newborns and children | Newborns and children aged < 37 weeks | Sanofi Pasteur | 2019 | (15) |
| Four-country Chronic Respiratory Disease (4CCORD) | To achieve consensus on a clinical diagnosis of chronic respiratory diseases (CRDs) (asthma, chronic obstructive pulmonary disease or 'other CRD') and identify the predictors of such diagnoses | Adults (18 years or over) | NIHR | 2019 | (16) |
| Health effects of living near unimproved brick kilns in Bangladesh | To estimate the impact of living near unimproved brick kilns on rates of pneumonia and other respiratory illnesses amongst children and adults | All age groups | Stanford University, USA | 2019 | (17) |
| Impact of 10-valent Pneumococcal Conjugate Vaccine (PCV-10) on Invasive Pneumococcal Disease (IPD) in Rural Bangladesh | To estimate the impact of PCV10 on laboratory-confirmed IPD and clinically identified severe pneumonia cases and its effectiveness on IPD caused by vaccine serotypes | Children aged 2-23 months | GlaxoSmithKline | 2019 | (7) |
| Determination of gestational age and preterm birth rates in low resource settings using newborn metabolic profiles | To predict the risk of preterm birth in low resource settings by measuring metabolites in maternal antenatal samples and neonatal blood samples taken at birth | Pregnant mothers and neonates | Stanford University, USA | 2020 | (18,19) |
| Campylobacter emergence and transmission in rural Bangladesh | To identify possible routes of Campylobacter transmission to young infants in households and communities | Pregnant women in their last trimester | George Mason University, USA | 2020 | In preparation |
| Real Time Tracking of Neglected Bacterial Diseases and Resistance Patterns in Asia (Tundra) | To measure the incidence of the respiratory syncytial virus (RSV) infection in the rural community of Bangladesh | Under-2 children | International Vaccine Institute (IVI), South Korea | 2023 | In preparation |
| Infection Bacterial Disease Surveillance | To provide information on invasive bacterial disease burden amongst under 5 children in Bangladesh | Under-5 children | WHO | 2023 | (20,21) |
| Metabolic Gestational Age Assessment | To predict the development of adverse health outcomes using maternal and neonatal metabolites | Pregnant women and newborns | Stanford University, USA | 2023 | In preparation |
| A Global Pediatric Cell Atlas of Nasal and Oral Mucosa | To characterize the cells present in nasopharyngeal cavity of children across the globe. The study is being undertaken in five countries in North America, Africa and Asia. Mirzapur is the collection site for Bangladesh arm of the study | Under-2 children | Boston Children's Hospital, USA (Chan Zuckerberg Initiative) | On going | - |
| Investigating variation in response to vaccines using single-cell sequencing | To understand the differences in immune response between preterm and term infants after vaccination | Term and preterm babies up to 40 weeks | Bill & Melinda Gates Foundation | On going | - |
| Development of capacity for neurodevelopmental assessment of children in Bangladesh for use in creating machine learning models for prediction of neurodevelopmental delays | To determine the metabolites responsible for neurodevelopmental delays | 3-4 years children | Stanford University, USA | On going | - |

The impact of pneumococcal conjugate vaccines (PCV10) was rigorously evaluated within the HDSS, showing a decrease in pneumonia-related hospitalizations among children under two (7). Additionally, research on nasopharyngeal pneumococcal carriage in vaccinated and unvaccinated cohorts provided valuable data on serotype shifts and vaccine effectiveness in South Asian settings (15).

The HDSS has also played a key role in maternal and newborn health research, including studies on Group B Streptococcus (GBS) colonization and its transmission from mother to newborn (13). Investigations into newborn metabolic profiles and gestational age assessments have advanced knowledge on preterm birth risks and neonatal health outcomes in resource-limited settings (18,19).

Studies conducted within the HDSS have highlighted the burden of chronic respiratory diseases, with findings from the 4CCORD survey revealing high rates of asthma in the region (16). Research on environmental health has demonstrated the adverse effects of air pollution from brick kilns, linking exposure to increased respiratory symptoms and deteriorating lung health among both children and adults (17).

Beyond infectious and respiratory diseases, the HDSS has supported global health initiatives, such as real-time tracking of neglected bacterial diseases and antimicrobial resistance patterns, as well as investigations into immune responses to vaccines at a cellular level (20,21). Ongoing research continues to explore the intersections between socio-economic factors, nutrition, and health outcomes in the Mirzapur community.

With its robust data infrastructure, longitudinal design, and strong community engagement, the Mirzapur HDSS remains a leading surveillance site for impactful public health research.

### Future Analysis Plan

The MHDSS will remain a dynamic platform, expanding to address emerging public-health needs:

1. Infectious and maternal-child health: evaluate long-term vaccine effectiveness, maternal immunization, antimicrobial-resistance trends and pediatric pathogens.
2. Non-communicable diseases and mental health: monitor diabetes, hypertension and cardiovascular risk factors alongside psychosocial well-being across age groups.
3. Environmental and climate impacts: assess how pollution, extreme weather and urbanization affect respiratory, infectious and other health outcomes.
4. Digital and multi-omics innovation: deploy AI models, digital tools and multi-omics for enhanced surveillance, prediction and early detection.
5. Healthcare access: study socio-economic, gender and geographic barriers to service use, with special focus on older adults.

### Main strengths and limitations

The MHDSS is a well-established surveillance system with high community engagement, real-time data collection, and strong research capacity. It has played a key role in generating high-quality longitudinal data that inform public health policies and interventions. One of the greatest strengths of the HDSS is its exceptionally high participation rate (99%), which is made possible through strong community trust and the involvement of locally trained data collectors, many of whom are women. This approach ensures accurate, reliable, and culturally sensitive data collection. Additionally, the integration of Android-based real-time data collection tools has significantly enhanced data accuracy, efficiency, and accessibility, making it a modernized and scalable model for demographic surveillance.

The integration of hospital and laboratory surveillance, particularly through the nested Infectious Disease Surveillance program, has strengthened the system’s ability to track infectious diseases and vaccine-preventable illnesses in children. MHDSS’s contribution to high-impact research on vaccine effectiveness, respiratory diseases, maternal and child health, and environmental health demonstrates its role as a valuable platform for global and national public health initiatives.

Despite its strengths, the HDSS faces challenges due to rapid urbanization and industrialization, which have increased population mobility and complexity in household tracking. To address this, the HDSS is exploring digital tracking tools and AI-driven monitoring to improve data collection efficiency. Additionally, seasonal flooding during monsoon months creates logistical obstacles for field teams, prompting efforts to incorporate remote data collection tools and satellite-based mapping to ensure data continuity. Finally, securing long-term funding remains a challenge, as maintaining large-scale, longitudinal surveillance systems requires sustained financial and technical support. Expanding international collaborations and diversifying funding sources will be critical for the HDSS’s continued success.

Despite these challenges, the Mirzapur HDSS remains a model for high-quality demographic surveillance, continuously evolving to address new public health challenges. By leveraging technology, strengthening collaborations, and adapting to emerging health trends, the HDSS is well-positioned to drive impactful research and inform evidence-based policies in Bangladesh and beyond.

### Can I get hold of the data? Where can I find out more?

Cohort data from the MHDSS are available upon request with a detailed description of the proposed project and approval from the CHRF Data Center. Requests can be submitted via email to.

## Ethical considerations

The study was approved by the Research and Ethical Review Committee of the Child Health Research Foundation.

## Author Contributions

SKS conceptualized the surveillance system; SKS and SS secured funding. SKS, SJ, GLD, SB, and SEA designed the surveillance framework. MSH, MSI, and ShS were responsible for implementation. NK, RCD, ATH, MSI and SAQ managed the data. SS, YH, NK and LT analyzed the data. SS, NK and LT drafted the manuscript with contributions from all authors. All authors reviewed and approved the final version. SS held the final responsibility for submitting the manuscript for publication.

## Use of Artificial intelligence (AI) tools

During the preparation of this work the authors used ChatGPT to check grammar, spellings, and improve readability of the manuscript. After using this tool, the authors reviewed and edited the content as needed and take full responsibility for the content of the publication.

## Acknowledgements

We would like to acknowledge Md. Mazharul Islam and the rest of the MHDSS field team member for their unwavering effort and contribution in maintaining the surveillance system. We appreciate the former and current community members of the MHDSS area for their cooperation. We also recognize our colleagues at KWMCH with gratitude for assisting us with hospital surveillance linked to the MHDSS.

## Data availability statement

The data underlying this article are available in the article and further data can be made available for collaborative research through CHRF’s Data Center upon request.

